# The impact of lung function on cardiovascular diseases and cardiovascular risk factors: a two sample bi-directional Mendelian randomization study

**DOI:** 10.1101/2020.06.01.20118919

**Authors:** SL Au Yeung, MC Borges, DA Lawlor, CM Schooling

## Abstract

**Background:** Observational studies suggested lung function is inversely associated with cardiovascular disease (CVD) although these studies could be susceptible to residual confounding. We conducted a 2 sample Mendelian randomization study using summary statistics from genome wide association studies (GWAS) to clarify the role of lung function in CVD and its risk factors, and conversely the role of CVD in lung function.

**Methods:** We obtained genetic instruments for forced expiratory volume in 1 second (FEV_1_) and forced vital capacity (FVC) from publicly available UK Biobank summary statistics (n = 421,986). We applied these genetic instruments for FEV_1_ (260) and FVC (320) to publicly available GWAS summary statistics for coronary artery disease (CAD) (n = 184,305), stroke and its subtypes (n = 446,696), atrial fibrillation (n = 1,030,836), and heart failure (n = 977,320) and cardiovascular risk factors. Inverse variance weighting was used to assess the impact of lung function on these outcomes. Sensitivity analyses included MR-Egger, weighted median, MR-PRESSO, and multivariable Mendelian randomization. We also conducted bi-directional Mendelian randomization to assess whether CVD affects lung function.

**Results:** FEV_1_ and FVC were inversely associated with CAD (odds ratio (OR) per standard deviation (SD) increase, 0.72 (95% confidence interval (CI) 0.63 to 0.82) and 0.70 (95%CI 0.62 to 0.78)), overall stroke (0.87 (95%CI 0.77 to 0.97), 0.90 (0.82 to 1.00)), ischemic stroke (0.87 (95%CI 0.77 to 0.99), 0.90 (95%CI 0.80 to 1.00)), small vessel stroke (0.78, (95%CI 0.61 to 1.00), 0.74 (95%CI 0.61 to 0.92)), and large artery stroke (0.69 (95%CI 0.54 to 0.89), 0.72 (95%CI 0.57 to 0.91)). FEV_1_ and FVC were inversely associated with type 2 diabetes (0.75 (95%CI 0.62 to 0.90), 0.67 (95%CI 0.58 to 0.79)) and systolic blood pressure. Sensitivity analyses produced similar direction for most outcomes although the magnitude sometimes differed. Adjusting for height attenuated results for CAD (e.g. OR for 1SD FEV_1_ 0.95 (0.76 to 1.20), but this may reflect weak instrument bias. This adjustment did not attenuate effects for stroke or type 2 diabetes. No strong evidence was observed for CVD affecting lung function.

**Conclusion:** Higher lung function likely protect against CAD and stroke.

## Introduction

Poorer lung function, indicated by reduced forced expiratory volume in 1 second (FEV_1_) and forced vital capacity (FVC), is associated with cardiovascular disease (CVD).^1–5^ However, these associations could be confounded by lifestyle factors, such as physical activity and smoking, and anthropometric characteristics, such as height, which are difficult to account for in observational studies.

Mendelian randomization is potentially less vulnerable to residual confounding than observational studies because it utilizes genetic variants related to exposures which are randomly allocated during conception.^6^ A previous two-sample Mendelian randomization study suggested higher FEV_1_ was causally related to lower risk of coronary artery disease (CAD), whilst the relation for FVC was less clear.^7^ However, that study used a relatively small number of genetic instruments (i.e.16 instruments for FEV_1_ (variance explained (R^2^): 0.8%) and 10 instruments for FVC (R^2^: 0.4%)), did not consider other important CVDs, such as stroke and heart failure, and did not explore the possibility of bidirectional effects (i.e. CVDs also having an effect on lung function).^7^ Furthermore, the genetic instruments used in that study had been adjusted for height and smoking and such adjustment can sometimes introduce bias in Mendelian randomization results.^8^ A more recent Mendelian randomization study, using UK Biobank and GWAS consortia data, concluded that most of the effect of height on CAD risk was mediated by lung function.^9^ However, that study did not examine the impact of lung function in detail (e.g. with detailed sensitivity analyses), its impact on other CVD outcomes, or possible reverse causation.

The aim of this study was to determine the causal effect of FEV_1_ and FVC on a wide range of cardiovascular diseases using 2 sample Mendelian randomization with summary statistics.^10^ Our study adds to previous Mendelian randomization studies by including more genetic instruments and more CVD outcomes and risk factors. We also explored whether genetic predisposition to CVDs might cause variation in lung function.

## Methods

### Study Design

We used two sample summary data Mendelian randomization to assess the effect of FEV_1_ (per standard deviation (SD)) and FVC (per SD) on multiple CVD outcomes: (i.e. CAD, stroke and its subtypes, heart failure, atrial fibrillation) and CVD risk factors (i.e. systolic and diastolic blood pressure, high density and low density lipoprotein cholesterol, triglycerides, type 2 diabetes, fasting glucose, glycated hemoglobin and insulin).^10^ Table 1 summarizes the study design including the sources of GWAS summary data for exposure (FEV1 and FVC – Sample 1) and each CVD outcome and risk factors (Sample 2). Given Mendelian randomization has stringent assumptions (i.e. relevance, independence and exclusion restriction), we also assessed whether the genetic instruments affected key confounders for CVD (education, body mass index (BMI), smoking, alcohol consumption, and height) to check exclusion restriction.^11^

**Table 1:** Data sources used in this Mendelian randomization study.

| **Exposure** | **Data source (PMID)** | **Sample size (% cases)** | **% European** |
| --- | --- | --- | --- |
| Forced expiratory volume in 1 second (SD) | UK Biobank (IEU GWAS) | 421,986 | 100 |
| Forced vital capacity (SD) | UK Biobank (IEU GWAS) | 421,986 | 100 |
| **Outcomes** | **Data source (PMID)** | **Sample size (% cases)** | **% European** |
| Coronary artery disease | CARDIoGRAM (26343387) | 184,305 (33%) | 77 |
| Stroke | MEGASTROKE (29531354) | 446,696 (9%) | 100 |
| Ischemic stroke | MEGASTROKE (29531354) | 440,328 (8%) | 100 |
| Cardioembolic stroke | MEGASTROKE (29531354) | 211,763 (3%) | 100 |
| Small vessel stroke | MEGASTROKE (29531354) | 198,048 (3%) | 100 |
| Large artery stroke | MEGASTROKE (29531354) | 150,765 (3%) | 100 |
| Atrial fibrillation | PMID: 30061737 | 1,030,836 (6%) | 100 |
| Heart failure | HERMES (31919418) | 977,320 (5%) | 100 |
| Systolic blood pressure (SD) | UK Biobank (IEU GWAS) | 436,419 | 100 |
| Diastolic blood pressure (SD) | UK Biobank (IEU GWAS) | 436,424 | 100 |
| LDL cholesterol (SD) | GLGC (24097068) | 173,082 | 100 |
| HDL cholesterol (SD) | GLGC (24097068) | 187,167 | 100 |
| Triglycerides (SD) | GLGC (24097068) | 177,861 | 100 |
| Glucose (mmol/L) | MAGIC (22581228) | 58,074 | 100 |
| HbA1c (%) | MAGIC (28898252) | 123,665 | 100 |
| Insulin (log) | MAGIC (22581228) | 51,750 | 100 |
| Type 2 diabetes | DIAGRAM (28566273) | 159,208 (17%) | 100 |
| **Confounders** | **Data source (PMID)** | **Sample size** | **% European** |
| Body mass index (SD) | GIANT (25673413) | 339,224 | 95 |
| Years education attained (SD) | SSGAC (27225129) | 328,917 | 100 |
| Alcohol (SD of Log transformed drinks per week) | GSCAN (30642351) | 941,280 | 100 |
| Smoking Heaviness (SD of cigarettes per day) | GSCAN (30642351) | 337,334 | 100 |
| Height (SD) | GIANT (25282103) | 253,288 | 100 |

### Ethics approval

In this study we have used publicly-available GWAS results from relevant publications and database (https://gwas.mrcieu.ac.uk/).^12–25^ No individual participant data were collected or used. Details of ethical approval and participant consent for each of the studies that contributed to the GWAS can be found in the original publications.

### Genetic determinants of FEV_1_ and FVC (Sample 1)

Genetic determinants of FEV_1_ (in standard deviation (SD), GWAS ID: ukb-b-19657) and FVC (SD, GWAS ID: ukb-b-7953) were extracted from summary genome-wide association study results in the UK Biobank, available in the IEU GWAS database https://gwas.mrcieu.ac.uk/)^16 17^ In brief, the UK Biobank is a large prospective cohort study where more than 500,000 participants were recruited in the United Kingdom (Great Britain only) from 2006 to 2010 (mean age: 56.5; 54% female). Pre-bronchodilation lung function testing was performed by trained healthcare staff using a Viitalograph Pneumotrac 6800 spirometer (Maids Moreton, UK). Genotyping was done using the Affymetrix UK BiLEVE Axiom array (∼50,000 participants) and Affymetrix UK Biobank Axiom array (∼450,000 participants). The summary statistics were generated using a linear mixed model to account for relatedness and population stratification, and adjusted for sex and genotyping array. The analyses were restricted to 421,986 participants of European descent. Quality controls included exclusion based on imputation quality (specific INFO scores based on minor allele frequency (MAF)) and MAF (< = 1%) (Supplemental Table 1).

### Genetic associations with the outcomes (Sample 2)

Complete summary GWAS results for CAD (CARDIoGRAMplusC4D 1000 Genome based GWAS),^12^ stroke and its subtypes (MEGASTROKE consortium),^13^ atrial fibrillation,^14^ heart failure (HERMES consortium),^15^ systolic and diastolic blood pressure (UK Biobank associations obtained from IEU GWAS database),^16 17^ LDL, HDL cholesterol and triglycerides (GLGC), ^18^ type 2 diabetes (DIAGRAM),^23^ fasting glucose, insulin, and glycated hemoglobin (MAGIC),^21 22^ were obtained from publicly available online GWAS summary data repositories, with majority of them retrieved via MR Base.^17^ Table 1 summaries the numbers (including the cases and controls where relevant) included in these GWAS, population (including ethnicity) and the sample size in the GWAS. Additional details, including quality control, imputation methods, and any covariates adjusted for in each GWAS and how the outcomes were defined are provided in Supplemental Table 1.

### Selecting genetic instruments from the exposure GWAS, identifying these in outcome GWAS and harmonizing instruments across studies

We identified genetic instruments for lung function at genome wide significant p value (< 5×10^8^), and we excluded instruments which were in high linkage disequilibrium (LD) with other instruments (r^2^< 0.001). We searched for the genetic instruments in the outcome datasets. For genetic instruments not available for an outcome, a proxy instrument in high LD with the original instrument (r^2^>0.8) was identified via MR-Base based on 1000 Genomes catalog (CEU reference population).^17^ No proxy instruments were identified for outcomes not available in MR-Base. We aligned each genetic association for exposure and outcome on the same effect allele. We used effect allele frequency to ensure palindromic genetic instruments were aligned properly where that was possible.

### Statistical analysis

We estimated the r^2^ of each genetic instrument and summed them up to compute the overall r^2^ and F statistics using the sample size which the instruments for lung function was derived from (n = 421,986). Higher r^2^ and F statistic values suggest lower risk of weak instrument bias.^26 27^ For our main Mendelian randomization analyses, we used inverse variance weighting (IVW) with multiplicative random effects to obtain the causal effect of FEV_1_ and FVC on CVD outcomes and their risk factors. IVW assumes there is no unbalanced horizontal pleiotropy.^28^ To assess the presence of potentially invalid instruments, we checked for evidence of heterogeneity across instrument-specific Wald ratios (i.e ratio of instrument-outcome association to instrument-exposure association) using Cochrane Q test. Heterogeneity may result from horizontal pleiotropy.

### Additional analyses to explore potential violation of Mendelian randomization assumptions

MR assumes that genetic instruments do not affect confounders of the exposure-outcome association. We assessed the plausibility of this assumption by exploring whether the FEV_1_ and FVC genetic instruments were related to key confounders defined as plausibly affecting lung function and CVDs (body mass index, height, socioeconomic position, alcohol and smoking).^11^ This was done using publicly available summary genome wide association study data for: body mass index,^20^ education (as a measure of socioeconomic position),^24^ number of drinks of alcohol per week and number of cigarette smoked per day,^19^ and height.^25^ Specifically, we meta analyzed the estimate of each SNP-outcome association (per lung function increasing allele) using standard inverse variance weighting method with additive random effects. For confounders which are associated with the genetic instruments, we then assessed whether these associations were a sign of vertical pleiotropy (causal role of lung function on confounders) or horizontal pleiotropy (causal role of confounders on lung function) using bi-directional Mendelian randomization.^11^ We used multivariable Mendelian randomization to control for potential horizontal pleiotropy by including estimates from the relevant confounders which were strongly associated with genetic instruments for lung function.^11 29^ We also approximated the conditional F statistics to evaluate potential weak instrument bias,^30^ presented Cochrane Q test to evaluate heterogeneity, and also provided estimates for multivariable MR-Egger to assess robustness of findings due to pleiotropy.^31^

To assess unbalanced horizontal pleiotropy bias we undertook the following sensitivity analyses: MR-Egger,^32^ weighted median analyses,^33^ and MR-PRESSO,^34^ that are more robust than IVW to the assumption that there is no effect of the genetic instrument on outcome other than through the exposures of interest (i.e. no horizontal pleiotropy paths).^28^ Like IVW, MR-Egger and MR-PRESSO require that the InSIDE (‘instrument strength is independent of direct effect’) assumption is not violated. Specifically, InSIDE assumes there is no correlation between the strength of the instrument (association of FEV_1_ or FVC on their respective genetic instruments) and the strength of any association of the genetic instruments on the outcome not via the exposures of interest (e.g. any horizontal pleiotropy paths). The weighted median analyses assumed at least 50% of the weight of the genetic instruments is via the exposure of interest. Further details of these methods and their additional (and differing) assumptions are available in the supplemental material.

### Bi-directional Mendelian randomization

To assess potential reverse causation, i.e. predisposition to CVDs affecting variation in FEV_1_ or FVC, we also conducted a Mendelian randomization using genetic predictors of risk of CAD, overall stroke, heart failure, and atrial fibrillation as instruments.^12–15^ We used IVW to explore evidence of effects of genetic predispositions to these CVDs on FEV_1_ or FVC.

All analyses were performed using R Version 3.5.2 (R Development Core Team, Vienna, Austria) using R packages (“TwoSampleMR”), and (“MRPRESSO).^17 34^

## Results

We included up to 260 SNPs for FEV_1_, which explained 3.5% of its variance and had an overall F statistic of 58.8. For FVC, we included up to 320 SNPs, which explained 4.8% of its variance and had an overall F statistic of 66.4. Supplemental Tables 2–3 shows the summary statistics of the lung function instruments used in this study.

Figure 1 shows the IVW MR causal effect estimates of FEV_1_ and FVC on the risk of CVD and type 2 diabetes. Higher FEV_1_ was associated with lower risk of CAD, overall stroke and some subtypes (i.e. ischemic stroke, small vessel stroke, large artery stroke), and type 2 diabetes, but increased risk of atrial fibrillation. FVC showed similar relation with these outcomes. There was no strong evidence that either FEV_1_ or FVC influenced heart failure (both point estimates were close to the null). Figure 2 shows the IVW MR causal effect estimates of FEV_1_ and FVC on cardiovascular risk factors using IVW. Higher FEV_1_ was mainly associated with lower systolic blood pressure and triglycerides. FVC showed similar relations with these outcomes and FVC was also associated with lower glucose and insulin. Supplemental Figures 1–4 show that between SNP Wald ratio heterogeneity was high for most of the outcomes.

**Figure 1:**
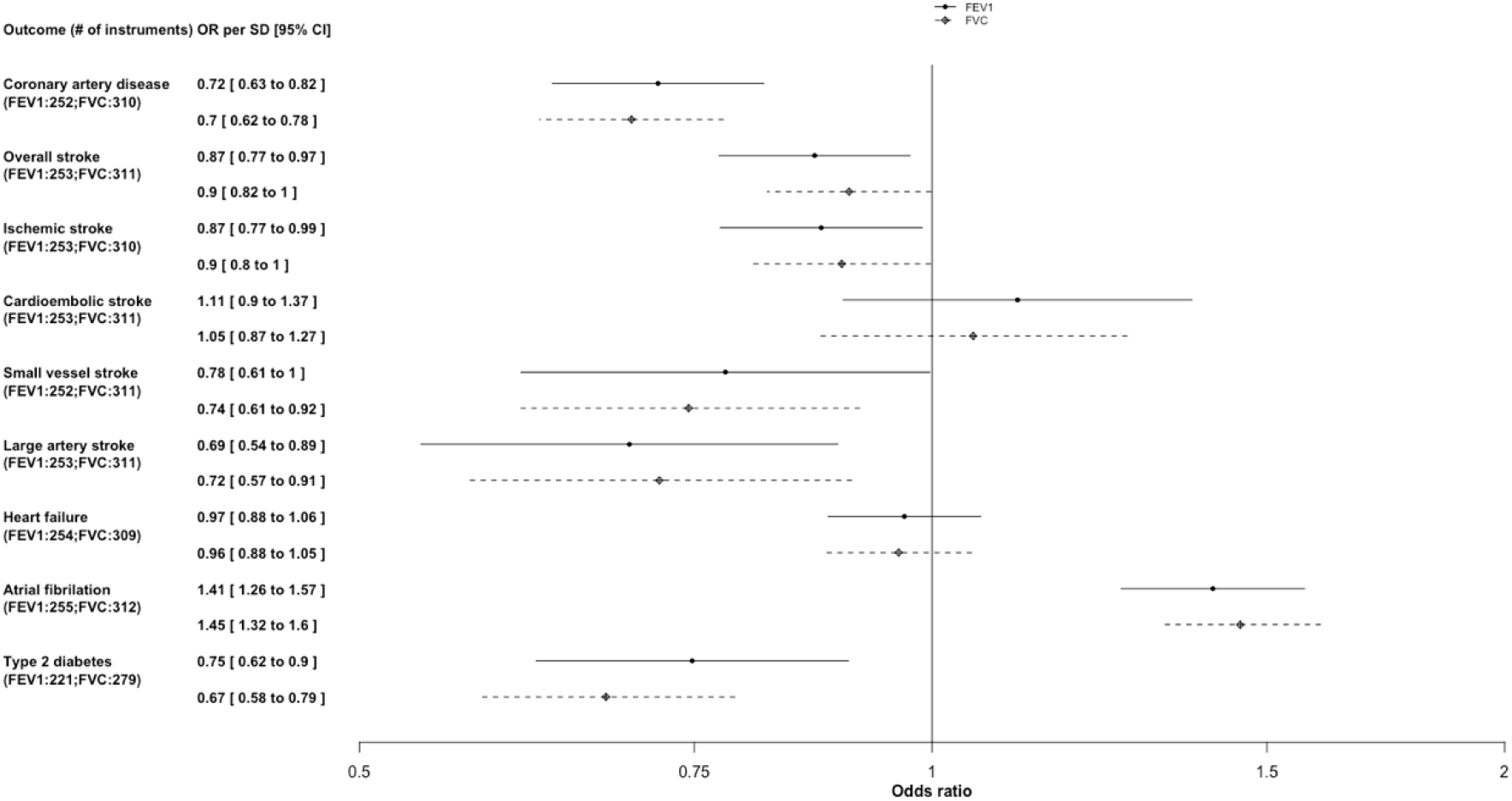
The impact of forced expiratory volume in 1 second (FEV_1_, per SD) and forced vital capacity (FVC, per SD) on cardiovascular disease and type 2 diabetes using Mendelian randomization.

**Figure 2:**
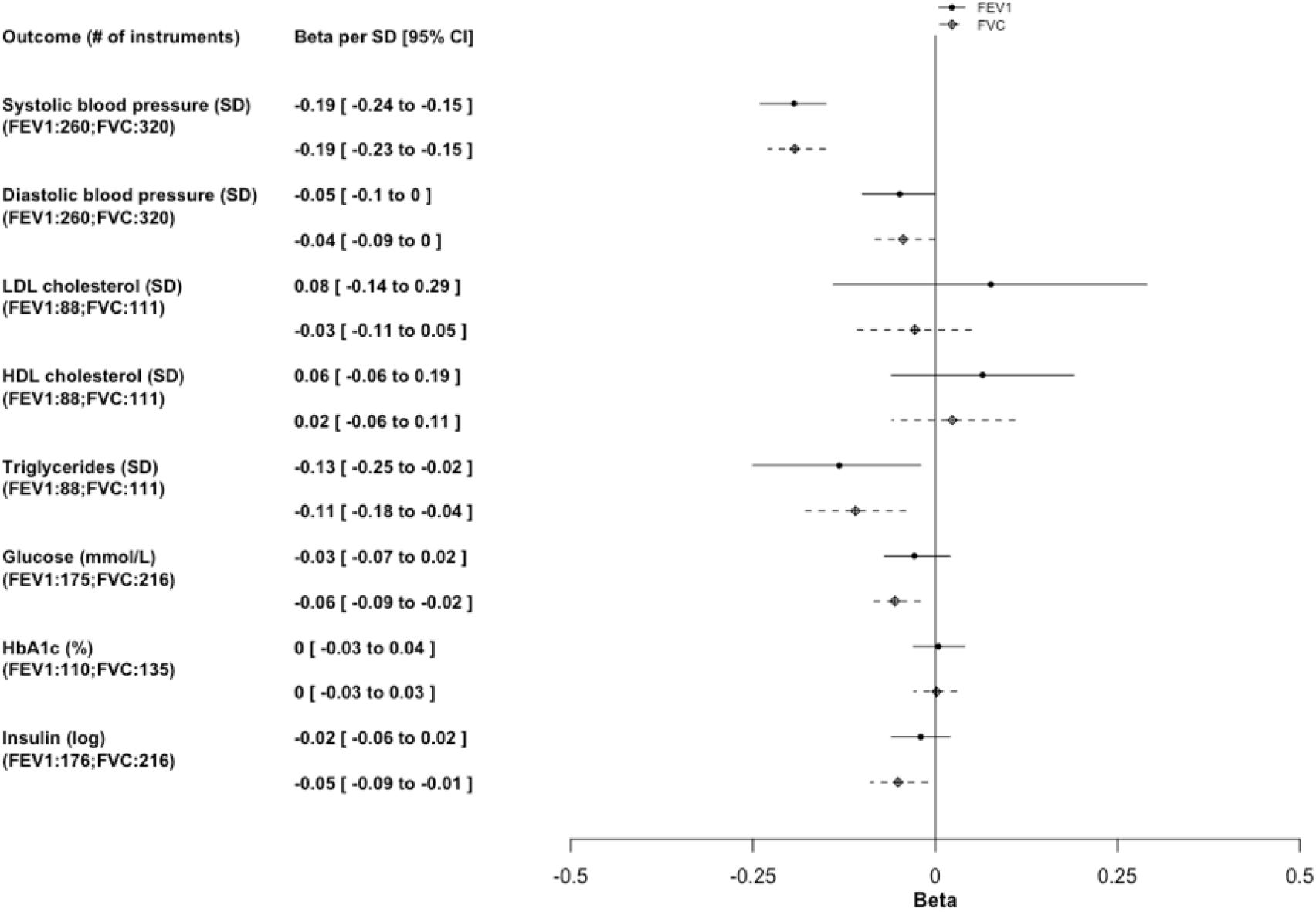
The impact of forced expiratory volume in 1 second (FEV_1_, per SD) and forced vital capacity (FVC, per SD) on continuously measured cardiovascular risk factors using Mendelian randomization.

**Figure 3:**
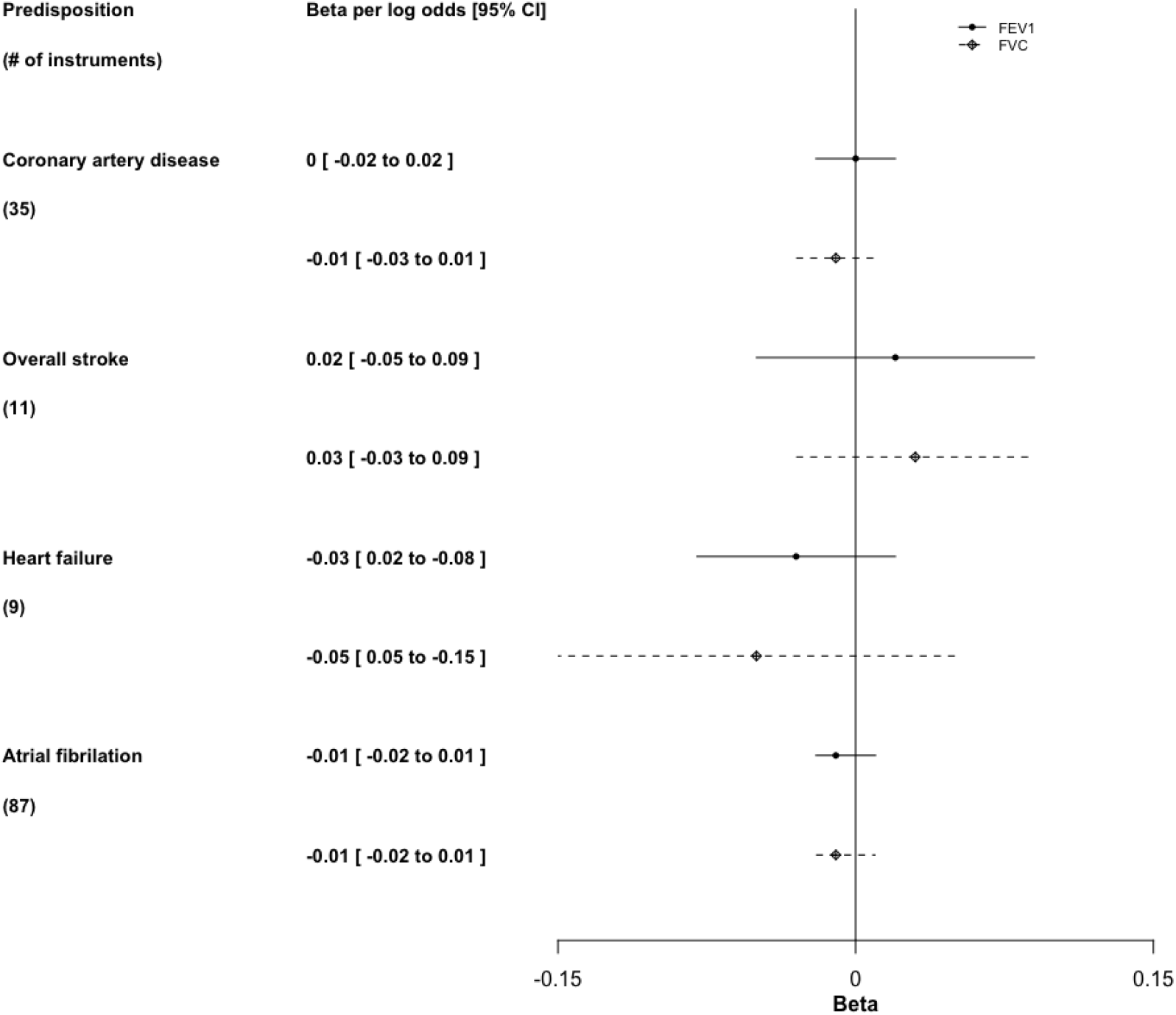
The impact of higher predisposition to cardiovascular outcome on forced expiratory volume in 1 second (FEV_1_) and forced vital capacity (FVC) using Mendelian randomization.

We found that genetic instruments for lung function were related to height, BMI, and education, with the strongest associations seen for height. There was no strong evidence of association with alcohol or tobacco phenotypes (Supplemental Table 4). Subsequent bi-Mendelian randomization analyses suggested there might be bidirectional effect between lung function and BMI, height, and education (Supplemental Table 5). Based on these findings we undertook multivariable MR adjusting for height only. This was done on the basis of the strong association of the lung function genetic instruments with height and knowledge that height is an established cause of lung function (Supplemental Table 4). As the associations of the genetic instruments for lung function with BMI and education were relatively weak and multivariable Mendelian randomization can introduce weak instrument bias we did not additionally adjust for these. Effect estimates from multivariable Mendelian randomization, adjusting for height, were directionally similar to the main IVW results. However, the results for CAD were attenuated quite notably (OR_MVMR-IVW_ for 1SD FEV_1_ 0.95 (0.75 to 1.19) and for 1SD FVC 0.87 (0.70 to 1.09), though height adjustment did not attenuate effects for stroke or type 2 diabetes (Supplemental Figures 5–8). With adjustment for height the positive association of FEV_1_ and FVC with atrial fibrillation disappeared whilst FEV_1_ and FVC were potentially associated with lower risk of heart failure (Supplemental Figures 5 and7). Similar findings were observed when we used multivariable MR-Egger method.

However, these analyses need to be treated with some caution as the conditional F statistics were small, ranging from 7.4 to 9.3 for FEV_1_ and 9.7 to 11.4 for FVC.

Supplemental figures 1–4 show the MR-Egger and weighted median effect estimates of FEV_1_ and FVC on CVD and cardiovascular risk factors. For FEV_1_, the direction and magnitudes of the estimates from MR-Egger and weighted median were consistent with those from IVW, with the exception of large artery stroke. The MREgger analysis suggested that higher FEV_1_ increased the risk of large artery stroke, albeit with wide confidence intervals overlapping null. However, the MR-Egger intercept test did not provide strong evidence for overall horizontal pleiotropy, except for atrial fibrillation. For FVC, the direction and magnitudes of the estimates from MR-Egger and weighted median were consistent with those from IVW. Similar findings to those seen from IVW analyses were observed when we used MRPRESSO to control for pleiotropy by outlier exclusion (Supplemental Tables 6–7).

Figure 3 shows the association of predisposition to cardiovascular disease with FEV_1_ and FVC, using genetic instruments for CVD (Supplemental Table 8). We found no strong evidence for an association between CVD predisposition and lung function.

## Discussion

In this Mendelian randomization study, we have assessed the impact of lung function on the risk of CVDs and risk factors which may mediate any observed effects on CVDs (blood pressure, lipids, glycemic traits). We also undertook several sensitivity analyses to explore assumption violations and explored possible confounding by height and smoking. We found convincing evidence that lung function is cardioprotective against CAD, consistent with our previous Mendelian randomization regarding the effect magnitude (OR_FEV1_: 0.78 and OR_FVC_: 0.82).^7^ These findings were consistent across a range of sensitivity analyses used to explore possible bias due to horizontal pleiotropy, however they did attenuate notably with multivariable Mendelian randomization adjustment for height (to an OR of 0.95 for FEV_1_ and 0.85 for FVC). We also found novel evidence that better lung function protects against stroke and type 2 diabetes, with these effects being consistent across sensitivity analyses, including adjustment for height. The positive association between lung function and atrial fibrillation in our main Mendelian randomization is in the opposite direction to that seen for other CVD outcomes but attenuated to the null with adjustment for height. The impact of lung function on heart failure is also less clear than that seen for CAD, stroke and type 2 diabetes, as the protective effect is only evident when adjusting for height in the multivariable Mendelian randomization. Lastly, our study also suggested that genetic susceptibility to CVD is unlikely to affect lung function.

Previous observational studies have suggested poorer FEV_1_ and FVC increase CVD.^1–5 35^ Our study suggests these observed associations are likely causal for CAD and stroke. Multiple pathways may explain the protective effect, with our study suggesting systolic blood pressure is a possible mechanism,^36^ but with weaker evidence of an effect of lung function on diastolic blood pressure. This may imply poorer lung function primarily acts on arterial stiffness.^37^ We also found better lung function associated with lower triglycerides, lower insulin and type 2 diabetes. As previous Mendelian randomization studies support a causal effect of hypertriglyceridemia, hyperglycemia, type 2 diabetes and higher insulin on CAD,^38–41^ these are possibly also mediators of the effect of lung function on CAD. With the exception of an inverse association with triglycerides, we did not find strong evidence for a causal effect of lung function on lipids. Importantly, we cannot rule out that the effect of lung function on CAD is largely explained by confounding due to height given the attenuation of the effect estimates in multivariable Mendelian randomization adjustment for height. These results have to be treated with some caution as the conditional F-statistics suggest possible weak instrument bias.

In our main analyses we found evidence of a protective effect of lung function on overall, ischemic, small vessel and large artery stroke, but not cardioembolic stroke. The underlying mechanisms may include reduced blood pressure,^42^ reduced risk of type 2 diabetes,^43^ whilst the role of lipid is not as clear.^41^ The lack of large GWAS with publicly available data on haemorrhagic stroke mean that we were not able to explore this outcome and it is not included in the overall stroke outcome. With the exception of MR Egger results sensitivity analyses were largely supportive of these causal effects. The magnitude of the effect estimates for these stroke outcomes were smaller than those for CAD and confidence intervals were wider, suggesting that lung function may be a weaker risk factor for stroke than CAD. However, it is also possible that the weaker effect of lung function on stroke (compared with that observed for CAD) is an underestimation because of selection bias. For example, if lung function is also related to survival then those who survive to have a stroke (which tend to occur at older ages than CAD) will likely be the ones with better lung function. Stroke shares several other risk factors, such as hypertension with CAD, but commonly occurs after CAD and this competing risk may attenuate or even reverse the estimates.^8 44^ Similar paradoxical findings have also been observed in observational studies, such as the association of cigarette smoking with dementia where the relation is attenuated, or even reversed with increasing age.^45^ Further replication of our findings for stroke in Mendelian randomization studies ideally in cohorts with younger age at recruitment may better help elucidate the role of lung function in stroke. In contrast to CAD and stroke, results for heart failure in the main analysis were very close to the null, which could suggest lung function is not causally related to heart failure. However, it remains possible that the lack of association is due to biases arising from covariable adjustments on survival and/or competing risk of CAD.^8 44^

The positive relation of lung function with atrial fibrillation in our main IVW analyses is somewhat unexpected given the majority of the findings from observational studies suggest an inverse relation.^46^ However, the positive association disappeared upon adjustment for height using multivariable Mendelian randomization. Whilst the reason underpinning these changes is unclear, it is possible that height may be a stronger confounder for atrial fibrillation than any other forms of CVD, or adjustment for height may have reduced biases arising competing risk of CAD. This could also imply substantial horizontal pleiotropy due to height could have made other pleiotropy robust methods (e.g. MR-Egger, MR-PRESSO) less efficient which gave the same findings with IVW in the atrial fibrillation analyses. Nevertheless, the results from multivariable Mendelian randomization should be interpreted with caution due to wider confidence intervals and the possibility of weak instrument bias.

Although we used Mendelian randomization which is less susceptible to confounding by key traits, such as smoking, height and socio-economic position,^47^ there are some limitations. First, the validity of our study depends on the 3 main instrumental variable assumptions.^6^ The F-statistics and proportion of variation in FEV_1_ and FVC make a major impact of weak instrument bias on our main IVW results unlikely. We chose SNPs identified from the UK Biobank summary statistics as instruments for lung function. Whilst this avoids the introduction of invalid SNPs due to bias arising from adjustment for height and smoking (as done in previous GWAS),^48^ the lack of replication of the UK Biobank GWAS results may mean that some of the genetic instrument-FEV_1_ and FVC associations are exaggerated and hence the Mendelian randomization results presented here attenuated. However, the broad similarity of our findings here for CAD with those from our previous paper (using GWAS summary data from a collaboration of studies not only including UK Biobank) suggest this has not caused major bias (OR_FEV1_: 0.78 and OR: 0.82_FVC_ in our previous study versus OR: 0.72_FEV1_ and OR: 0.70_FVC_ in this study).^7^ We also explored whether the lung function genetic instruments were associated with potential confounders of the relation of lung function with CVDs risk (i.e. BMI, smoking, alcohol and education, and height), which if present could bias our results through horizontal pleiotropy. We found limited evidence that the lung function instruments relate strongly to these risk factors, apart from height. Multivariable Mendelian randomization to adjust for height did notably attenuate the effects on CAD, though not those for stroke or type 2 diabetes. However, additional exploratory analyses suggested the magnitude, but not direction, of some multivariable Mendelian randomization estimates were sensitive to the choice of GWAS where height instruments were extracted from, such as directly from GIANT (Supplemental Figures 5–8) or UK Biobank (Supplemental Figures 9–12). As such, interpretation of these adjusted results is difficult because of their imprecision and the possibility of weak instrument bias and it would be valuable to repeat our analyses using data from even larger summary GWAS. Whilst we found evidence for between SNP heterogeneity, which might occur because of horizontal pleiotropy resulting in violation of the third assumption, sensitivity analyses that explore bias due to unbalanced horizontal pleiotropy (i.e. MR-Egger, weighted median and MR-PRESSO analyses) supported our main analyses. In addition to these three key assumptions, two-sample MR assumes that the two samples are from the same underlying population and that the two samples are independent. With the exception of summary data for CAD (23% non-European), all of our samples were restricted to European populations. There was overlap between the lung function and blood pressure (100% – i.e. both were undertaken in UK biobank), atrial fibrillation (38% of this outcome GWAS participants were in UK biobank), and heart failure (40% in UK biobank). Such overlap can mean that any weak instrument bias could be towards confounded estimates, rather than towards the null (as in two sample MR with independent samples). This would particularly influence blood pressure and could mean associations are exaggerated by in the presence of weak instruments. Second, we were unable to explore the possibility of non-linear effects between lung function and CVD, which can only be explored in one sample Mendelian randomization in large biobanks with individual level data.^49^

In conclusion, our Mendelian randomization study provides some evidence concerning the protective role of lung function on CAD, stroke, type 2 diabetes and lower systolic blood pressure. Future studies should explore the underlying mechanisms, including the role of height in these relationships, and hence help identify additional targets of intervention for cardiovascular disease prevention.

Future GWAS and Mendelian randomization studies exploring causes of hemorrhagic stroke are also important, particularly for settings where hemorrhagic stroke is prevalent, such as China.^50^

## Data Availability

All data used in this study are publicly available and can be accessed via the references and links presented in the manuscript.

## Acknowledgement

Data on coronary artery disease have been contributed by CARDIoGRAMplusC4D investigators and have been downloaded from www.CARDIORAMPLUSC4D.ORG. We thank the investigators of the atrial fibrillation GWAS for providing the summary statistics, which have been downloaded from http://csg.sph.umich.edu/willer/public/afib2018/. We thank MEGASTROKE for providing the summary statistics for stroke, which have been downloaded from http://www.megastroke.org/index.html. The MEGASTROKE project received funding from sources specified at http://www.megastroke.org/acknowledgments.html. The list of authors in the MEGASTROKE Consortium can be found in the Supplemental materials. We thank HERMES Consortium for providing the summary statistics for heart failure, which have been downloaded from http://www.broadcvdi.org/informational/data. We thank GLGC for providing summary statistics for lipids, which have been downloaded from http://csg.sph.umich.edu/willer/public/lipids2013/. We thank MAGIC for providing summary statistics for glycemic traits, which have been downloaded from https://www.magicinvestigators.org. We thank DIAGRAM for providing the summary statistics for type 2 diabetes, which have been downloaded from https://www.diagram-consortium.org. We thank GIANT consortium for providing the summary statistics for body mass index and height, which have been downloaded from https://portals.broadinstitute.org/collaboration/giant/index.php/GIANT_consortium_data_files. We thank SSGAC for providing the summary statistics for years of education attainment, which have been downloaded from https://www.thessgac.org/data. We thank GSCAN for providing the summary statistics for alcohol and cigarette use phenotypes, which have been downloaded from https://conservancy.umn.edu/handle/11299/201564. Data on FEV_1_ and FVC, systolic and diastolic blood pressure were extracted from summary genome-wide association study results in the UK Biobank, available in the IEU GWAS database (https://gwas.mrcieu.ac.uk/). We thank Mr Wong KN for summarizing the descriptive information regarding the GWAS used in this study.

## Authors’ contribution

SLAY designed the study, wrote the analysis plan and interpreted the results. SLAY undertook analyses with feedback from MCB, DAL, and CMS. SLAY wrote the first draft of the manuscript with critical feedback and revisions from MCB, DAL, and CMS. All authors gave final approval of the version to be published. SLAY had primary responsibility for final content.

